# Meconnaissance in Headache Disorders: Does the ICHD3 Inherently Allows for Misrecognition in Cases of Multiple Headaches?

**DOI:** 10.1101/2025.03.02.25323173

**Authors:** Pengfei Zhang

## Abstract

**Background:** In clinical circumstances with more than one headache disorders, misrecognition can occur if each headache disorder is not separately and individually evaluated. For example, consider the case of a patient with baseline tension type headaches lasting hours who now develops cluster headaches with nausea. Merging the characteristics of both disorders may yield the illusion of migraine. Do these types of misrecognitions occur necessarily because of ICHD3 criteria? Are there combinations of ICHD3 criteria, if any, which necessarily yield this type of misrecognition?

**Objective:** This work seeks to determine whether new, unintended diagnosis, can be arrived when one merges two ICHD3 diagnoses.

**Methods:** We translated ICHD3 primary headache disorders, as well as headache phenotypes closely mimicking ICHD3 criteria, as numerical data using prime encoding method. We then model the encoding of two concurrent diagnoses in an individual by multiplying all pairing of ICHD3 encoding. We then diagnose the result algorithmically to see which pairings, if any, generate “new”/” illusionary” diagnoses.

**Results:** A total of 117,526 phenotypes were generated. After duplicate pairing of diagnoses were removed, we derived a total of 72 pairings of unique ICHD3 diagnoses that yield a “new” diagnosis. Thirty-seven (51%) of these pairings were not judged to be “illusionary”: they simply arise from our model’s idiosyncrasy of encoding chronic migraine without encoding migraine with or without aura explicitly. Of the rest, 23 pairings involve combining paroxysmal hemicrania with either a longer or shorter duration headache.

We further generalize the condition under which the above “illusionary” diagnosis occurs as a mathematical observation.

**Conclusion:** Misrecognition arises naturally if diagnostic criteria consist solely of characteristics from another headache disorder or a combination of two headache disorders. Specifically, combining headaches with different duration and/or frequency are most likely to yield these “illusionary” diagnoses.

## Background

In clinical circumstances with more than one headache disorder, misdiagnoses could occur if each disorder is not individually evaluated. Consider, for example, the scenario where a patient has baseline tension type headaches. Should this patient develop new cluster headache with nausea, the combination of exhibited symptoms - headache lasting 30 minutes to 7 days (from baseline tension type headaches), unilaterality (from new cluster headaches), and association with nausea - may suggest to the clinician a misdiagnosis of migraine.^1^

Could these kinds of misrecognitions arise naturally as result of how we define headache disorders in the International Classification of Headache Disorders (ICHD3)? In other words, are there pairings of headache disorders, when occurring simultaneously, that would lead to an “illusionary” third diagnosis simply by the way we design diagnostic criteria?

The result of this question carries significant implications for clinical diagnoses and research. For the former, if misrecognitions are inevitable whenever two specific disorders arise concurrently, then an increase likelihood for misdiagnosis would be built-in to our classification system. These misdiagnoses may, in turn, lead to errors in inclusion/exclusion of patients in clinical trials.

This work seeks to provide the answer for these questions of misrecognition in ICHD3. Specifically, we enumerate and merge all possible pairings of diagnoses from the ICHD3. We then determine which, if any, of these pairings yields a third unintended diagnosis. Finally, we seek to determine the condition of possibility of misrecognition; our goal is to determine the rules which make this sort of misrecognition possible from a classification point of view, thereby providing a framework to identify and/or avoid them in future classifications.

## Methods

To study classification criteria *en masse*, one needs to translate ICHD3 into manipulable numerical data. In a prior paper, we demonstrated that it is possible to do so through the following^2^: First, we represent every headache characteristic (i.e. photophobia, phonophobia, nausea, etc…) as a prime number. We then represent every headache phenotype within a headache diagnosis as composite numbers by the following algorithm:

1. We first translate headache criteria into a series of logic AND statements connected by OR statements. (This is called “disjunctive normal form” and can be done for all Boolean logic statements.)
2. For every AND that is used between two characteristics, the corresponding prime numbers for those two characteristics are multiplied together. The results are called encodings.
3. All composite numbers that are generated, and separated by an OR statement, are then combined into a list.

The above is feasible due to the connection between logical statements and prime number composition. (Indeed, our encoding method is merely a variant of Gödel numbering.^3^) We will briefly describe the above algorithm through an example but refer the interested reader to the original article for more information. The complete list of encoding as well as prime number representation are also presented in Table 1 and 2 of our prior article.^2^For the ease of the reader, prime representations are reproduced in Appendix A of this text with permission.

Consider the ICHD3 criteria for cluster headache, we can represent all possible phenotypes of disorder as the following series of composite numbers.

1577982193416431, 1932584933510011, 2251727399594233, 3492836989921763, 4237502744118281, 4911247950296083, 7428927404960501, **7960831515100871**, 8173593159157019

Similarly, for infrequent tension type headaches:

1378253359942681, 1386197183342927, 2206388426002661, 2219105362175587, 2241879928833803, **2254801426982701**, 9699783080351321, 9755689611073807, 9855811762608983, 9912617594093761, 15777758744434123, 15868696835180141, 514088503258620013, 517051549386911771, 522358023418276099, 525368732486969333, 836221213455008519, 841040932264547473, 3676217787453150659, 3697406362596972853, 194839542735016984927, 195962537217639561209 Important to our discussion, the encoding algorithm contains the ability for automated diagnosis through modular arithmetic: when the phenotype of a specific patient’s headache profile, encoded by prime number, can be divided by the encoding of a diagnosis without remainder, then that diagnosis is satisfied for the patient according to the ICHD3.

For example, consider a patient with both infrequent tension type headache and episodic cluster headaches, the autonomic feature being restlessness, with symptoms of nausea. The set of characteristics, as well as its associated prime encoding, is the following according to the ICHD3: nausea (281), > 5 episodes (157), severe (479), unilateral (523), orbital or supraorbital (383), duration between 15 minutes to 3 hours (11), restlessness (449), one every other day to 8 per day (107), > 10 episodes of headaches (257),< 12 days per year (199), 30 minutes to 7 days (19), bilateral (53), not aggravated by routine activities (379), no nausea (331), no photophobia (349), and no phonophobia (347).

(Notice that in this specific case the patient would describe “no nausea” for his tension type headaches but with nausea for his new onset of cluster.)

Multiplying the above sequences of prime encoding would generate the following composite number:

1750259841031105929758798277984900497

Notice that this number is divisible by 7960831515100871 and 2254801426982701 without remainder. The former of these two numbers is an encoding for the criteria of cluster headaches whereas the latter is an encoding for tension type headaches. (These are bolded in the encodings above.) Therefore, this patient has a diagnosis of cluster headache and infrequent tension type headache. This example is not an isolated occurrence; rather it can be proven that given a set of headache characteristics, the above algorithm would work for automated diagnosis of any ICHD3 headache disorders.

### Modification to the encoding to account for implications

Notice that the above encoding cannot be divisible by any of the encodings for migraine without aura and so it is not a diagnosis for the above combinations of characteristics. The reason is that although a headache that lasts “30 minutes to 7 days” (19) could also be a headache that lasts between “4 hours to 72 hours” (23), a migraine criterion, the two characteristics are encoded separately in the original database.

To account for these kinds of implications, we derived a list of characteristics which *implies or could imply* the other in Table 1. For example, the criterion “15 minutes to 180 minutes” (11) necessarily implies the criterion “30 minutes to 7 days in duration” (19), since any duration that satisfies the former must also satisfy the latter. (In other words, the former criterion is a subset of the latter.) We therefore include this in Table 1. On the other hand, a headache that lasts 170 minutes can be thought of as having the duration of hours, therefore the criterion “15 minutes to 180 minutes” (11) is also an implication of “hours to days” (179). (In other words, there exists an element in the former criterion which is also in the latter criterion.) The decision to include the latter stems from the need to capture and model ambiguity in clinical circumstances such as those presented in our motivating example.

### Merging diagnoses

Once Table 1 is generated, any encoding that contains a characteristic in column 1 of Table 1 is then multiplied by the implied characteristics of column 2. This artificially adds the implied criterion to the original. The result is a numerical database of clinical circumstances representing all possible phenotypes of headache diagnoses as well as close mimics of each characteristic.

Given this list, we then calculate the combination of any pairing of two headache phenotypes by multiplying each phenotypical encoding with another phenotypical encoding. (Similar to our numerical example above.) Keeping track of which diagnoses we are merging, we then applied our aforementioned automated diagnosis technique (through the unmodified, original, ICHD3 encoding) to the multiplied list.

The above algorithms are generated and implemented through custom codes in Haskell.

## Results

Twenty-seven logical implication statements were obtained (see Table 1) and the encoding database was modified accordingly.

Using the new encoding data, a total of 117,526 phenotypes were generated through merging pairings of headache phenotypes. Each was then diagnosed through automated diagnosis. Duplicates are possible in our methodology because multiple different kinds of pairing of phenotypes are possible for any two given headache disorders. (For example, various different phenotypes of migraine without aura may be combined with the various phenotypes of cluster headaches.)

Table 2 represents a complete table of 72 pairings in which a new “illusionary” diagnosis is generated. Notice that in the above results, some of the diagnoses are not actually “illusionary” but rather contain artifacts from our model’s idiosyncrasy of encoding chronic migraine: in the ICHD3, chronic migraine is required to be either migraine with aura or migraine without aura. As a result, the encoding for chronic migraine always includes within it encoding for migraine with aura or without aura. Therefore, whenever chronic migraine is merged with another diagnosis, inevitably the result would include an additional diagnosis of migraine without aura or migraine with aura. There are 37 of such results, or 51.1% of the 72 pairings. Those that are not caused by this idiosyncrasy are highlighted with an asterisk on Table 2.

Two pairings in our result yield a 5 concurrent headache diagnoses. Both of these arise from the combination of paroxysmal hemicrania and chronic migraine. Part of the reason is from the idiosyncrasy of chronic migraine encoding as discussed above. However, combining paroxysmal hemicrania and chronic migraine can still produce the illusion of cluster and SUN. The reason for this is two-fold: 1. Since SUN is defined as between 1 to 600 seconds and PH between 2 minutes to 30 minutes, there is an overlap in timeframe of these diagnoses. Since SUN is not defined as indomethacin non-responsive, this allows for concurrent diagnoses of both. 2. PH and cluster headache duration also overlap. As a result, when PH is merged with chronic migraine criteria, a headache that is at least 15 days per month, the criteria of frequency for cluster headache is also satisfied, allowing for a diagnosis of the latter. In other words, CM criteria lead to the illusion of a longer and more frequent headaches of TAC phenotype, distorting the original diagnosis that can lead to two more potential headaches.

Indeed, PH, when merging with another headache of different duration and frequency, allows for distortion and therefore the illusion of a new diagnosis can be seen in 23 of pairings in Table 2. Specifically, combining PH with a shorter lasting headache – such as primary sex headaches, cough headaches, primary stabbing headaches, exercise headaches, traction headache – lead to the illusionary diagnosis of SUN. This is logical since one can interpret the short duration to be associated with the TAC features. Combining PH with a longer lasting headache – such as new daily persistent headache – leads to the illusion of hemicrania continua. This pattern of “time-distortion” also goes the opposite direction: combining a long-lasting headache - such as chronic tension type headache or HC - with a very short-lasting headache – such as SUN – produce medium duration headache such as cluster or PH as an illusion.

### Generalization

It would be beneficial to produce a general rule in constructing classification guidelines that would allow us to avoid illusionary pairings. That is, we can determine the condition of possibility in which illusionary diagnosis occurs. This is possible by thinking of diagnostic criteria as their prime number encodings; we present the following theorem as well as its mathematical proof.

### Theorem

Given two different, modified, diagnoses encodings, *p* and *q*. Let *d* be the encoding of an illusionary diagnosis that arises from the combination of both. Then *d* exists (i.e. *d* does not equal *p* nor *q*) only if its characteristics are some combination of the characteristics of *p* and *q* or their subsets.

### Proof

Let *p* = *p*_*1*_*p*_*2*_…*p*_*i*_…*p*_*n*_and *q = q*_*1*_*q*_*2*_*…q*_*i*_…*<H* unique prime encoding of characteristics. Assume that *d* exists, then it must divide *p* multiply by *q* without remainder. Then *d* must divide some combination of *p*_*i*_and *q*_*i*_without remainder. Since all *p*_*i*_and *q*_*i*_are prime numbers and, by definition, *d* does not equal *p* nor *q*, then *d* must either: 1. Divide *p* and therefore is the product of some subset of *p*_*i*_. 2. Divide *q* and therefore is the product of some subset *q*_*i*_, 3. Equal the multiplication of some combination of *p*_*i*_and *q*_*i*_.

QED

The proof above arises naturally from our construction of prime number encoding as well as its ability to automate diagnoses. Furthermore, it suggests that we can avoid illusionary diagnosis so long as any new headache disorder– that is, *d*, in our proof – does not contain solely the characteristics, nor related characteristics such as those in Table 1, within other pairings of headache diagnoses.

## Discussion

This paper evaluates how two concurrent headache diagnoses may be combined to create a new one. We also showed, mathematically, the theoretical condition under which these “illusionary” diagnoses can be generated/avoided. On a practical level, a key result is that merging headaches with divergent frequency and/or duration are the most susceptible to generating misrecognition. This is particularly true if headache triggers – such as cough, sexual activities, etc. – of the so called “other primary headache disorders” are ignored. On a theoretical level, another key result is that any new diagnostic criteria the characteristics of which can be made a subset, or a combination of subsets, of pre-existing diagnostic criteria will allow for misrecognition to naturally occur.

While the above results have clear implications for the clinical practice of headache medicine and the creation of future iterations of diagnostic criteria, we believe the implication of this work is not limited to classification. Indeed, classification should not be thought of as rules, rather they are crystallization of our intuition of headache diagnoses in general. What then do the possibilities of illusionary diagnosis in the ICHD3 tell us about our current intuition regarding headache disorders?

Unconventionally, this discussion will take two opposing perspectives on this phenomenon. 1. Splitters: we will first discuss how from a classification perspective, illusionary diagnoses should be unwanted. 2.

Lumpers: we will then discuss the possibility of the opposite, where illusionary diagnosis may actually be the key to more precise diagnoses.

### Splitters

That, when encountering different kinds of headaches, one should address each headache individually, is a paradigm common in headaches. Most recently, this approach is well-articulated in an influential article by Deborah Friedman.^4^From this perspective, the existence of illusionary headaches highlights the importance of detailed history taking. Indeed, it is a specific kind of detailed history taking that is required: in order to root out illusionary diagnoses, not only are clinicians required to pay attention to details, but they are also required to guide the patient in self-discernment. That is, a skilled clinician ought to be able to help the patient communicate and reflectively separate divergent headache phenotypes. Our paper shows that this is most important in the diagnoses of TACs, where duration are often the key features, and as a result, the misrecognition of divergent coexisting primary headaches may disrupt and distort the final diagnoses.

While our paper limits itself to primary headaches, the inclusion of secondary headaches further complicates misrecognition. In real life settings, primary headaches exist alongside secondary headaches, the latter often having a wide variety of duration and/or frequency. In the case of CSF leak, for example, the onset/resolution of headaches varies widely after position change.^6^As shown in our paper, co-existence of headaches of different duration may lead to misrecognition and generation of illusionary diagnoses.

Therefore, the need for clinicians to dissect and be able to discern the existence of secondary headaches on top of primary headaches in a complicated patient presentation is then even more important.

Of crucial importance is the existence of medication overuse headaches in this regard. MOH has been reported to change primary headache phenotype.^7^This therefore complicates the diagnostic process by placing headache phenotype in a dynamic/changing setting. Furthermore, the existence of MOH is itself premise on existence of two headaches. The situation quickly becomes complicated in patients who have baseline primary headache disorders who then suffered a common secondary headache - such as post-traumatic headache – and subsequently self-medicate. In these circumstances, the number of headache disorders one needs to clarify may increase rather quickly.

Our result does appear to offer an incomplete solution for the clinicians in these settings: in the case of primary headaches, and in particular TACs, misrecognition appears to occur frequently with primary headaches of short durations with known triggers. For example, the misrecognition of SUN in the setting of a combination of primary sex headaches, cough headaches, cold induced headaches, in the setting of PH. Therefore, although often rare and “exotic”, the identification of the existence of “other primary headache disorders” together with perhaps the incongruent duration of a patient’s headache on history, may hints to a careful clinician the existence of multiple disorders.

In the light of our generalized result, the above is not an isolated incident that belongs exclusively to triggers. Indeed, the reason that triggers can play an important role is because they are very specific and contain pathognomonic characteristics that differentiate one diagnosis from all else. In other words, the reason that some headache diagnoses are easily merged is because they have characteristics that are not pathognomonic. For example, most of the problems arise from duration, which can be shared amongst many kinds of headaches.

### Lumpers

“*Psycho-analysis regards the consciousness as irremediably limited, and institutes it as a principle, not only of idealization, but of Meconnaissance, as – using a term that takes on new value by being referred to a visible domain – scotoma*.” (Jacques Lacan, The Four Fundamental Concepts of Psychoanalysis)^5^

Does the patient know? Most certainly some patients don’t know or at least cannot tell – the demented patient, the autistic patient, the patient with severe post-traumatic disorder, the patient with psychosis. But who else also does not know?

The foundation of clinical medicine is history taking. No specialty, aside from perhaps psychiatry, is more heavily reliant on history taking than headache medicine. Yet, history relies on the assumptions that the patient knows his or her diseases. In our field, for example, one assumes that the patient possesses the ability and the know-how, subjectively, to separate different headaches. In the very least, when prompted with questions of different headaches, they are supposed to be able to know the differences even if they themselves may not know the precise diagnosis.

This paper’s results challenge that assumption implicitly. What if often the patient does not know? Or, in a Lacanian way, they misrecognize a single presentation as separate or multiple as the same?

In circumstances where a secondary headache occurred after a baseline primary headache, the differentiation may be easy. For example, a head injury which produces a new posttraumatic headache in someone with a baseline history of migraine.

However, when two primary headaches coexist, are these really two different headaches or is it simply one headache that the patient misinterprets? For example, when someone tells us a story of having primary stabbing headache in the presence of hemicrania continua, does that make the so-called illusionary new diagnosis of SUN an actual misdiagnosis? Or perhaps it is really the patient who is misleading us to believe there are two separate disorders when they simply have SUN? In this perspective, the real diagnosis is the illusionary – since it is, by the famous Occam’s razor, the most unifying one. Perhaps we, as diagnosticians, are the ones who have been misled along with the patient who themselves has what Lacan would call *Meconnaissance*.

The clinical implication here from this perspective is clear: perhaps lamotrigine actually would be the drug of choice in the above example as opposed to indomethacin.

### Limitations

Our paper is not without limitations. One of the first limitations is that the dataset is limited to primary headaches. Future direction of this work would be the inclusion of both secondary headaches as well as facial pain. Furthermore, our encoding system does not make determinations for the clinician for the existence or lack of existence of each characteristic but rather defer that to the user. For example, our model does not determine whether a patient should be represented as having nausea when he has that symptom in 1 out of 10 headaches. The decision to encode that is left to the clinician. Therefore, our automated diagnosis system, while can be implemented in real life, does require clinicians as careful historians.

## Conclusion

Our result shows that there are certain restrictions to our intuitions of headache diagnoses as classified by the ICHD3. Namely, the existence of *Meconnaissance* when two headaches co-exist shows that our conception of specific combinations of headaches – specifically those of trigeminal autonomic cephalalgias – may require further refinement. Rather than siding with the “lumpers” or the “splitters”, in clinical circumstances these opposite perspective may hold the keys to open up possibilities of alternative interpretation of clinical presentation rather than closing them.

### Article highlights

- Combining two different headaches with differences in duration and frequency may cause the illusion of a third unrelated headache disorder.
- If a new disorder has characteristics which are the subsets of combinations of characteristics from two other headache disorders, then that new disorder can become an “illusionary” disorder.

## Data Availability

All data produced in the present study are available upon reasonable request to the authors

## Glossary

ICHD3: International classification of headache disorder, 3^rd^Edition
SUN: short lasting unilateral neuralgiform headaches
CM: Chronic migraine
MwA: migraine with aura
NDPH: new daily persistent headache
HC: hemicrania continua
PH: paroxysmal hemicrania
MOH: medication overuse headache
PTH: posttraumatic headache

## Declaration of Interests

PZ: He has received honorarium from Alder Biopharmaceuticals, Board Vitals, and Fieve Clinical Research. He collaborates with Headache Science Incorporated without receiving financial support. He had ownership interest in Cymbeline LLC.

### Study Funding

Beth Israel Deaconess Medical Center/Harvard Medical School provides open access fees for publication of this article.

## Acknowledgements

None

## References

1. The International Classification of Headache Disorders, 3rd edition. Cephalalgia 2018;38:1–211

2. Zhang P. Which headache disorders can be diagnosed concurrently? An analysis of ICHD3 criteria using prime encoding system. Front Neurol. 2023 Aug 21;14:1221209. doi: 10.3389/fneur.2023.1221209. Erratum in: Front Neurol. 2024 Apr 08;15:1404283. doi: 10.3389/fneur.2024.1404283. PMID: 37670775; PMCID: PMC10475541.

3. Gödel K, “Über formal unentscheidbare Sätze der Principia Mathematica und verwandter Systeme I”, Monatshefte für Mathematik und Physik, 38: 173–198, doi:10.1007/BF01700692

4. Friedman DI. Approach to the Patient With Headache. Continuum (Minneap Minn). 2024 Apr 1;30(2):296–324. doi: 10.1212/CON.0000000000001413. PMID: 38568485.

5. Lacan, J. (1977). The Four Fundamental Concepts of Psycho-Analysis (J.A. Miller, Ed.) (1st ed.). Routledge. 10.4324/9780429481826

6. Mehta D, Cheema S, Glover S, Qureshi AM, Davagnanam I, Kamourieh S, Sayal P, Toma A, Lagrata S, Joy C, Duncan C, Anderson J, Davies B, Dorman PJ, Angus-Leppan H, Walkden J, Rohrer J, Matharu MS. Defining the typical characteristics of orthostatic headache in patients with spontaneous intracranial hypotension. Cephalalgia. 2025 Jan;45(1):3331024241308154. doi: 10.1177/03331024241308154. PMID: 39781568.

7. Diener, HC., Kropp, P., Dresler, T. et al. Management of medication overuse (MO) and medication overuse headache (MOH) S1 guideline. Neurol. Res. Pract. 4, 37 (2022). 10.1186/s42466-022-00200-0

