## Supplementary material for "Meconnaissance in Headache Disorders: Does the ICHD3 Inherently Allows for Misrecognition in Cases of Multiple Headaches?": Table 1

| Table 1 |  |
| --- | --- |
| Column 1 | Column 2 |
| 1 min to 24 hours with severe pain (2) | 1 to 600 seconds (5), 15minutes up to 4 hours after waking (7), 15 to 180 minutes (11), 2 to 30 minutes (17), 30 min to 7 days (19), 21 to 14 days per month (3), 1 second to 2 hours (47), hours to days (179), less than 48 hours (211), severe (479),greater than 5 minutes (163) |
| 1 to 14 days per month (3) | more than 10 days per month (269) |
| 1 to 600 seconds (5) | 1 min to 24 hours with severe pain (2), 2 to 30 minutes (17), 1 second to 2 hours (47), less than 48 hours (211), up to a few seconds (557),greater than 5 minutes (163) |
| 15minutes up to 4 hours after waking (7) | 1 min to 24 hours with severe pain (2), 15 to 180 minutes (11), 2 to 30 minutes (17), 30 min to 7 days (19), 1 second to 2 hours (47), less than 48 hours (211), up to 72 hours with mild intensity (547),greater than 5 minutes (163) |
| 15 to 180 minutes (11) | 1 min to 24 hours with severe pain (2), 15minutes up to 4 hours after waking (7), 2 to 30 minutes (17), 30 min to 7 days (19), 1 second to 2 hours (47), less than 48 hours (211), up to 72 hours with mild intensity (547),greater than 5 minutes (163) |
| 2 to 30 minutes (17) | 1 min to 24 hours with severe pain (2), 1 to 600 seconds (5), 15 to 180 minutes (11), less than 48 hours (211), up to a few seconds (557),greater than 5 minutes (163) |
| 30 min to 7 days (19) | 1 min to 24 hours with severe pain (2), 15minutes up to 4 hours after waking (7), 15 to 180 minutes (11), 4 to 72 hours (23), 1 second to 2 hours (47), less than 48 hours (211), up to 72 hours with mild intensity (547),greater than 5 minutes (163) |
| 4 to 72 hours (23) | 1 min to 24 hours with severe pain (2), 30 min to 7 days (19), less than 48 hours (211), up to 72 hours with mild intensity (547),greater than 5 minutes (163) |
| 1 second to 2 hours (47) | 1 min to 24 hours with severe pain (2), 1 to 600 seconds (5), 15minutes up to 4 hours after waking (7), 15 to 180 minutes (11), 2 to 30 minutes (17), 30 min to 7 days (19), less than 48 hours (211), up to 72 hours with mild intensity (547), up to a few seconds (557),greater than 5 minutes (163) |
| greater than 1 per day (137) | greater than 15 days per month (139), greater than 5 per day (167), greater than 8 days per month (173), more than 1 episode per day (251), more than 10 episodes (257),more than 10 days per month (269) |
| greater than 15 days per month (139) | every other day to 8 per day (107),more than 10 days per month (269) |
| greater than 2 episodes (149) | greater than 1 per day (137), greater than 20 episodes (151), greater than 5 episodes (157),more than 10 episodes (257) |
| greater than 20 episodes (151) | greater than 1 per day (137), greater than 2 episodes (149), greater than 5 episodes (157),more than 10 episodes (257) |
| greater than 5 episodes (157) | greater than 1 per day (137), greater than 2 episodes (149), greater than 20 episodes (151),more than 10 episodes (257) |
| greater than 5 minutes (163) | 1 min to 24 hours with severe pain (2), 1 to 600 seconds (5), 15minutes up to 4 hours after waking (7), 15 to 180 minutes (11), 2 to 30 minutes (17), 30 min to 7 days (19), 4 to 72 hours (23), 1 second to 2 hours (47), less than 48 hours (211),up to 72 hours with mild intensity (547) |
| greater than 5 per day (167) | greater than 1 per day (137),more than 1 episode per day (251) |
| greater than 8 days per month (173) | 1 to 14 days per month (3), every other day to 8 per day (107), greater than 15 days per month (139),more than 10 days per month (269) |
| hours to days (179) | 1 min to 24 hours with severe pain (2), 15minutes up to 4 hours after waking (7), 15 to 180 minutes (11), 30 min to 7 days (19), 4 to 72 hours (23),1 second to 2 hours (47) |
| less than 48 hours (211) | 1 min to 24 hours with severe pain (2), 1 to 600 seconds (5), 15minutes up to 4 hours after waking (7), 15 to 180 minutes (11), 2 to 30 minutes (17), 30 min to 7 days (19), 4 to 72 hours (23), 1 second to 2 hours (47), greater than 5 minutes (163), hours to days (179), up to 72 hours with mild intensity (547), up to a few seconds (557) |
| mild to moderate pain (233) | 1 min to 24 hours with severe pain (2), moderate to severe (241),up to 72 hours with mild intensity (547) |
| moderate to severe (241) | 1 min to 24 hours with severe pain (2), mild to moderate pain (233), severe (479) |
| more than 1 episode per day (251) | every other day to 8 per day (107), greater than 1 per day (137), greater than 15 days per month (139), greater than 1 per day (137),more than 10 days per month (269) |
| more than 10 episodes (257) | greater than 1 per day (137), greater than 2 episodes (149), greater than 20 episodes (151),greater than 5 episodes (157) |
| more than 10 days per month (269) | 1 to 14 days per month (3), greater than 15 days per month (139), greater than 20 episodes (151),greater than 8 days per month (173) |
| severe (479) | 1 min to 24 hours with severe pain (2),moderate to severe pain (241) |
| up to 72 hours with mild intensity (547) | 1 to 600 seconds (5), 15minutes up to 4 hours after waking (7), 15 to 180 minutes (11), 2 to 30 minutes (17), 4 to 72 hours (23), 1 second to 2 hours (47), less than 48 hours (211),mild to moderate pain (233) |
| up to a few seconds (557) | 1 to 600 seconds (5), 1 second to 2 hours (47),less than 48 hours |
