## Supplementary material for "Meconnaissance in Headache Disorders: Does the ICHD3 Inherently Allows for Misrecognition in Cases of Multiple Headaches?": Table 2

| **Table 2** |  |  |  |  |  |  |  |
| --- | --- | --- | --- | --- | --- | --- | --- |
| **Headache 1** | **Headache 2** | **A result of CM encoding?** | **Diagnosed Headache 1** | **Diagnosed Headache 2** | **Diagnosed Headache 3** | **Diagnosed Headache 4** | **Diagnosed Headache 5** |
| cm | chronic tension type headache | * | migraine w/o aura | cm | frequent tension type headache | chronic tension type headache |  |
| cm | chronic tension type headache |  | mwa | cm | chronic tension type headache |  |  |
| cm | chronic tension type headache | * | mwa | cm | frequent tension type headache | chronic tension type headache |  |
| cm | cluster |  | cluster | mwa | cm |  |  |
| cm | cluster |  | migraine w/o aura | cluster | cm |  |  |
| coldHA | cm |  | coldHA | mwa | cm |  |  |
| coldHA | cm |  | migraine w/o aura | coldHA | cm |  |  |
| compression | cm |  | compression | mwa | cm |  |  |
| compression | cm |  | migraine w/o aura | compression | cm |  |  |
| cough | cluster | * | cluster | sun | cough |  |  |
| cough | cm |  | migraine w/o aura | cm | cough |  |  |
| cough | cm |  | mwa | cm | cough |  |  |
| exercise | cluster | * | exercise | cluster | sun |  |  |
| exercise | cm |  | exercise | mwa | cm |  |  |
| exercise | cm |  | migraine w/o aura | exercise | cm |  |  |
| frequent tension type headache | cm |  | migraine w/o aura | cm | frequent tension type headache |  |  |
| frequent tension type headache | cm |  | mwa | cm | frequent tension type headache |  |  |
| hc | cm |  | hc | mwa | cm |  |  |
| hc | cm |  | migraine w/o aura | hc | cm |  |  |
| hypnic | chronic tension type headache | * | hypnic | frequent tension type headache | chronic tension type headache |  |  |
| hypnic | cm |  | migraine w/o aura | cm | hypnic |  |  |
| hypnic | cm |  | mwa | cm | hypnic |  |  |
| infrequent tension type headache | cm |  | migraine w/o aura | cm | infrequent tension type headache | frequent tension type headache |  |
| infrequent tension type headache | cm |  | mwa | cm | infrequent tension type headache | frequent tension type headache |  |
| infrequent tension type headache | hypnic | * | hypnic | infrequent tension type headache | frequent tension type headache |  |  |
| migraine w/o aura | cm |  | migraine w/o aura | mwa | cm |  |  |
| migraine w/o aura | hypnic | * | migraine w/o aura | cm | hypnic |  |  |
| mwa | cm |  | migraine w/o aura | mwa | cm |  |  |
| mwa | hypnic | * | mwa | cm | hypnic |  |  |
| ndph | cm |  | migraine w/o aura | ndph | cm |  |  |
| ndph | cm |  | ndph | mwa | cm |  |  |
| nummular | cm |  | migraine w/o aura | cm | nummular |  |  |
| nummular | cm |  | mwa | cm | nummular |  |  |
| ph | chronic tension type headache | * | cluster | ph | chronic tension type headache |  |  |
| ph | chronic tension type headache | * | cluster | ph | sun | chronic tension type headache |  |
| ph | cluster | * | cluster | ph | sun |  |  |
| ph | cm | * | cluster | ph | mwa | cm |  |
| ph | cm | * | cluster | ph | sun | mwa | cm |
| ph | cm | * | migraine w/o aura | cluster | ph | cm |  |
| ph | cm | * | migraine w/o aura | cluster | ph | sun | cm |
| ph | coldHA | * | coldHA | ph | sun |  |  |
| ph | compression | * | compression | ph | sun |  |  |
| ph | cough | * | ph | sun | cough |  |  |
| ph | exercise | * | exercise | ph | sun |  |  |
| ph | frequent tension type headache | * | ph | sun | frequent tension type headache |  |  |
| ph | hc | * | ph | sun | hc |  |  |
| ph | hypnic | * | ph | sun | hypnic |  |  |
| ph | infrequent tension type headache | * | ph | sun | infrequent tension type headache |  |  |
| ph | migraine w/o aura | * | migraine w/o aura | ph | sun |  |  |
| ph | mwa | * | ph | sun | mwa |  |  |
| ph | ndph | * | ndph | ph | hc |  |  |
| ph | ndph | * | ndph | ph | sun | hc |  |
| ph | nummular | * | ph | sun | nummular |  |  |
| sex | cluster | * | cluster | sun | sex |  |  |
| sex | cm |  | migraine w/o aura | cm | sex |  |  |
| sex | cm |  | mwa | cm | sex |  |  |
| sex | ph | * | ph | sun | sex |  |  |
| stabbing | cluster | * | cluster | sun | stabbing |  |  |
| stabbing | cm |  | migraine w/o aura | cm | stabbing |  |  |
| stabbing | cm |  | mwa | cm | stabbing |  |  |
| stabbing | ph | * | ph | sun | stabbing |  |  |
| sun | chronic tension type headache | * | cluster | sun | chronic tension type headache |  |  |
| sun | cm |  | migraine w/o aura | sun | cm |  |  |
| sun | cm |  | sun | mwa | cm |  |  |
| sun | hc | * | ph | sun | hc |  |  |
| thunderclap | cluster | * | thunderclap | cluster | sun |  |  |
| thunderclap | cm |  | migraine w/o aura | thunderclap | cm |  |  |
| thunderclap | cm |  | thunderclap | mwa | cm |  |  |
| thunderclap | ph | * | thunderclap | ph | sun |  |  |
| traction | cm |  | migraine w/o aura | traction | cm |  |  |
| traction | cm |  | traction | mwa | cm |  |  |
| traction | ph | * | traction | ph | sun |  |  |
